# Personality correlates of past-year alcohol use in individuals with severe alcohol use disorder and a lifetime history of involvement in Alcoholics Anonymous

**DOI:** 10.1101/2024.02.16.24302941

**Authors:** Séverine Lannoy, Dace S. Svikis, Mallory Stephenson, Kathryn Polak, Kenneth S. Kendler, Alexis C. Edwards

## Abstract

**Background:** Alcohol Use Disorder (AUD) is a highly impairing condition with important public health impacts. Despite the availability of treatment options for AUD, research shows that few people receive treatment, and even fewer can maintain abstinence/low drinking levels. This study investigated the role of personality traits in current alcohol use among individuals with severe AUD who ever attended Alcoholics Anonymous (AA), a widespread and easily accessible self-help group for alcohol problems.

**Methods:** Univariable and multivariable regressions were performed separately in females and males with alcohol consumption as an outcome. Socioeconomic factors, genetic liability, and psychopathology were included as covariates.

**Results:** Results from the multivariable model indicated that in females who attended AA, higher alcohol use was related to both positive and negative urgency as well as low sensation seeking, while in males, higher alcohol use was related to positive urgency. Results also indicated an important role of younger age and lower educational levels in higher alcohol use in both sexes. Moreover, single males and those with lower AUD severity were at higher risk of using alcohol in the past year.

**Conclusions:** These findings highlighted sex-specific correlates of drinking in individuals with AUD who engaged in self-help groups. Findings may be useful to improve treatment options, as personality encompasses modifiable traits that can be targeted in psychological interventions.

## Introduction

Alcohol Use Disorder (AUD) is an important public health burden related to widespread diseases, disabilities, and premature mortalities (<u>World Health Organization, 2021</u>). AUD is defined by DSM-5 criteria referring to physiological, psychological, and social consequences of drinking (<u>DSM-5, 2013</u>), which further subdivide AUD by levels of severity (mild AUD: 2-3 symptoms, moderate AUD: 4-5 symptoms, and severe AUD: more than 6 symptoms). AUD is a quite common disorder with a past-year and lifetime prevalence of, respectively, 14% and 30% in US samples (<u>Grant et al., 2015</u>), yet this disorder is characterized by low treatment adherence and success (<u>Grant et al., 2017</u>). To reduce the personal and societal burdens of AUD, research should focus on improving our understanding of treatment-related outcomes.

Over 60 years of research have facilitated significant advances in AUD treatment, offering three complementary options (<u>NIAAA, 2014</u>): medication, behavioral treatments (e.g., cognitive and behavioral therapy), and mutual-support groups (e.g., Alcoholics Anonymous). Alcoholics Anonymous (AA) is a focus of interest in the scientific literature. AA is a worldwide peer-to-peer support group with a 12-step program (<u>Kelly et al., 2020</u>). There is mixed evidence about the effectiveness of AA (<u>Kaskutas, 2009</u>): Some studies indicate that it is related to worse outcomes than no treatment (<u>Kownacki and Shadish, 1999</u>), others show that it has no empirical effectiveness (<u>Ferri et al., 2006</u>), and still others find that it is causally associated with abstinence (<u>Humphreys et al., 2014</u>). The most recent systematic reviews and meta-analyses generally support a positive association between participation in AA and abstinence (e.g., <u>Kelly et al., 2020</u>, <u>Adriana and Wicaksono, 2023</u>). Nevertheless, there is a dearth of knowledge about the specific factors associated with positive outcomes in individuals attending AA recovery groups. Prior studies indicate that the achievement of abstinence is related to improvement in self-efficacy, psychosocial well-being, and the development of new social networks (<u>Adelman-Mullally et al., 2021</u>, <u>Pfund et al., 2023</u>), but less is known about personality characteristics that may describe successful AA participants and, specifically, how these personality traits are related to alcohol use.

AUD is a heterogeneous syndrome (<u>Litt et al., 1992</u>, <u>Babor et al., 1992</u>, <u>Cloninger et al., 1996</u>, <u>Cloninger et al., 1981</u>), with different subtypes characterized by differences in age of onset, biological sex, genetic liability, personality, and psychiatric comorbidity (<u>Babor, 1996</u>, <u>Leggio et al., 2009</u>, <u>Kendler et al., 2022</u>). AUD is highly comorbid with certain personality disorders (<u>Newton-Howes and Foulds, 2018</u>), and personality traits related to impulsivity (novelty seeking and low persistence) have been found to distinguish individuals who maintained abstinence from those who continued to drink alcohol in clinical trials (<u>Foulds et al., 2017</u>). However, discrepancies exist regarding the role of personality traits in treatment outcomes (<u>Newton-Howes et al., 2017</u>). These discrepancies might be explained by the effects of confounders: (i) personality traits differ according to sex (<u>Schmitt et al., 2008</u>), while sex also influences response to AUD treatment (i.e., heavy alcohol drinking after treatment is more common in males) (<u>Sofin et al., 2017</u>); (ii) personality traits are partially heritable (<u>Sanchez-Roige et al., 2018</u>) and genetically correlated with alcohol use and problems (<u>Sanchez-Roige et al., 2019</u>). Shared genetic liability might thus account for the co-occurrence of impulsive personality traits and heavy drinking; and (iii) additional factors related to alcohol drinking and/or heavy alcohol drinking after treatment need to be controlled for, such as AUD symptoms/severity (<u>Kirouac and Witkiewitz, 2019</u>, <u>Martins et al., 2022</u>), comorbid depression (<u>Neighbors et al., 2005</u>), criminal/antisocial behaviors, and sociodemographic factors, i.e., younger age, male sex, being single, being unemployed, having lower educational attainment (<u>Sofin et al., 2017</u>).

The current study explored associations between personality and recent alcohol use in individuals with severe AUD who reported attending AA. To overcome previous limitations, we conducted sex-specific analyses, considered the influence of genetic liability for alcohol problems, and controlled for a range of sociodemographic variables, psychopathology, and clinical features. Our goal was to provide insight into whether features of personality are related to greater success – operationalized as lower alcohol consumption in the past year – among AA attendees. We evaluated the roles of personality measures using two validated self-report instruments: facets of the Big Five (extraversion, agreeableness, conscientiousness, neuroticism) and UPPS-P (negative and positive urgency, lack of premeditation, lack of perseverance, sensation seeking). After accounting for sociodemographic variables, psychopathology, and AUD severity, we expected the association between personality traits and higher past-year alcohol use in individuals who attended AA to differ by sex (<u>Schmitt et al., 2008</u>, <u>Foulds et al., 2017</u>), with traits such as low perseverance, low conscientiousness, and high sensation seeking more prominent in males, and emotional traits such as negative urgency and neuroticism more evident among females. Providing a better understanding of who is at risk of continuing to drink heavily after participation in AUD recovery groups is useful because it might help refine treatment approaches. Moreover, findings could inform the design of additional behavioral interventions (e.g., neuropsychological training may help reduce impulsivity and improve response to AUD treatment).

## Materials and Methods

### Sample

Participants were from the Genes, Addiction, and Personality (GAP) study, a cross-sectional research study that investigates genetic and environmental influences on severe AUD. Participant recruitment was organized (i) in person through substance use disorder treatment facilities and (ii) online through recovery community organizations, Facebook advertisements, and ResearchMatch.org. Participants completed the survey either using a custom-designed app (<u>Ondersma et al., 2011</u>) for in-person recruitment or REDCap (<u>Harris et al., 2009</u>, <u>Harris et al., 2019</u>) for online recruitment. The current analyses were limited to 2,966 individuals from the online recruitment (65.8% female; mean age = 47.28 years, range = 18-79 years), who met criteria for a lifetime history of severe AUD (6+ symptoms), and reported a history of involvement in AA. To define involvement in AA, participants were asked “In your lifetime, which of the following types of addiction treatment programs or services have you received?” where “AA or other self-help groups” was a response option. Participants provided written consent and study procedures were approved by VCU Institutional Review Board.

### Measures

#### Outcome

We used the Alcohol Use Disorders Identification Test-Consumption (<u>AUDIT-C; Bush et al., 1998</u>) to evaluate past-year alcohol drinking. AUDIT-C measures alcohol consumption frequency, intensity, and the number of drinking episodes in which participants drank more than six alcohol units on one occasion. As some participants in our sample were actively involved in treatment, we added a response option to the AUDIT-C, i.e., “In treatment/recovery and not currently using alcohol”. This response has been recoded as 0 (not drinking) for the current analysis. We used the sum score of the three items as the dependent measure (range: 0-12).

#### Personality

First, we used the Big Five Inventory, which is designed to measure the most common personality traits (<u>Hayes and Joseph, 2003</u>). In this study, we included items evaluating four of these traits: extraversion, agreeableness, conscientiousness, and neuroticism. Openness was not included in the assessment due to prior evidence demonstrating minimal correlation with psychopathology, including substance use disorders (<u>Kotov et al., 2010</u>), and to minimize participant burden. Items were rated on a five-point Likert scale from 1 (disagree a lot) to 5 (agree a lot) and we computed subscores for each dimension (range 0-30 for the neuroticism trait and 0-15 for other personality traits), with a higher score reflecting a higher tendency for a specific trait.

Second, impulsivity was measured using 15 items from the short version of the UPPS-P impulsive behavior scale (<u>Cyders et al., 2014</u>). This instrument included three items for each dimension of impulsivity: positive and negative urgency, defined as the tendency to act rashly in response to positive and negative emotions; lack of premeditation, which is the tendency to not consider the long-term consequences of an action; lack of perseverance, defined by the difficulty to complete a boring or difficult task; and sensation-seeking, which is the tendency to seek excitement and new sensations. Responses were recorded on a 4-point Likert scale, from 1 (disagree strongly) to 4 (agree strongly). We created a mean score for each of the four subscales (range 1-4), with higher scores reflecting higher impulsivity.

#### Sociodemographic variables

We considered four sociodemographic covariates: age, education, self-reported race, and relationship status. Education level was measured with eight levels collapsed into four categories for the current analysis: High school or less, some post-secondary education, Bachelor degree (set as reference), and Graduate degree. Due to relatively low sample sizes in many self-reported race categories, we also collapsed race into 3 groups: White or Caucasian (reference group), Black or African American, and Other which included Asian, American Indian or Alaskan Native, Native Hawaiian, or Other Pacific Islander. Finally, we included information on whether individuals were married/cohabited (reference group) with someone or divorced/separated/living alone.

#### Family history density

We constructed a family history risk score by calculating a weighted sum of the number of family members with a history of alcohol problems (i.e., fathers, mothers, full siblings, sons, and daughters were weighted at 0.50; half-siblings, grandfathers, grandmothers, aunts, and uncles were weighted at 0.25) divided by the total family size (<u>Stephenson et al., 2022</u>). We standardized the score (mean = 0 and SD = 1) for the whole GAP sample before selecting our study sample (online participants who reported AA participation).

#### AUD severity

The number of symptoms was counted from the DSM-5 definition of AUD (<u>DSM-5, 2013</u>) based on items referring to the participant’s heaviest drinking period. All participants included in the current study endorsed at least 6 of the criteria.

#### Comorbid psychopathology

We considered lifetime depression using a narrow definition of major depressive disorder related to the worst depressive episode (<u>DSM-5, 2013</u>), corresponding to endorsement of a core symptom (i.e., sadness or loss of interest); at least 5 symptoms total; symptoms’ persistence for at least two weeks; and symptoms’ interference with daily functioning.

Adult antisocial behavior was measured using 17 items evaluating the occurrence of antisocial behaviors since turning 18 (<u>see Stephenson et al., 2022 for more details</u>). We used a sum score, with higher values reflecting a higher occurrence of antisocial behaviors.

### Statistical analysis

Descriptive statistics were computed for the study sample. Univariable and multivariable linear regressions were performed separately for males and females with AUDIT-C as an outcome. The main predictors included personality traits: extraversion, agreeableness, conscientiousness, and neuroticism as well as negative and positive urgency, lack of premeditation, lack of perseverance, and sensation seeking. All analyses included age, race, family history of alcohol problems, relationship status, education, AUD severity, major depression, and antisocial behavior as covariates.. Analyses were performed using R 4.2.1 (<u>R Core Development Team, 2021</u>) and the lm function. Missing data were handled with listwise deletion; therefore, only individuals with complete data for all predictors and covariates contributed to the final models (see Supplement for information on missing data). In addition to the sex-specific approach, we pursued supplementary analyses to formally evaluate whether sex-by-personality-trait interactions were associated with AUDIT-C (Supplement).

## Results

### Descriptive analyses

The sample included 1953 females and 1016 males. Table 1 provides a full description of the sample. In females, the mean AUDIT-C was 2.85 (SD=3.57) and 38% of the sample had a score higher than 3, indicating harmful alcohol use (<u>Saunders et al., 1993</u>). In males, the mean AUDIT-C was 3.68 (SD=4.10), with 43% of the sample having a score higher than 4 indicating harmful drinking.

**Table 1.** Descriptive statistics

| Variables: Mean (SD) or n |  |  |
| --- | --- | --- |
|  | Females, N=1953 | Males, N=1013 |
| Age | 45.76 (12.74) | 50.25 (13.54) |
| Race |  |  |
| White | 1762 | 879 |
| Black | 79 | 56 |
| Other | 107 | 47 |
| In a relationship | 1021 | 482 |
| Education |  |  |
| High school or less | 245 | 154 |
| Post-secondary education | 842 | 447 |
| Bachelor degree | 508 | 264 |
| Graduate degree | 357 | 196 |
| AUD severity | 11.5 (2.28) | 11.83 (2.27) |
| Major depression (n) | 1545 | 670 |
| Family history | 0.25 (0.98) | 0.06 (1.05) |
| Positive urgency | 2.12 (0.82) | 2.20 (0.83) |
| Negative urgency | 2.74 (0.81) | 2.62 (0.82) |
| Lack of perseverance | 1.93 (0.66) | 1.92 (0.67) |
| Lack of premeditation | 2.19 (0.73) | 2.06 (0.69) |
| Sensation seeking | 2.45 (0.79) | 2.69 (0.77) |
| Conscientiousness | 12.49 (2.37) | 12.39 (2.45) |
| Extraversion | 9.80 (3.41) | 9.35 (3.40) |
| Neuroticism | 20.30 (4.04) | 18.71 (4.25) |
| Agreeableness | 11.89 (2.27) | 11.39 (2.45) |
| Antisocial behaviors | 16.36 (10.60) | 17.85 (10.59) |

Figure S1 presents the correlations between the study variables (Supplement).

### Linear regressions

Results from univariable regression models are presented in Table 2 and multivariable models in Table 3. Associations between each covariate and AUDIT-C score are available in Table S1 (Supplement). Univariable regressions for personality traits were controlled for sociodemographic factors, genetic liability, and psychopathology, and showed that most personality traits were associated with higher alcohol use in both males and females, including high positive and negative urgency, lack of perseverance, lack of premeditation, low conscientiousness, high neuroticism, and low agreeableness. Only extraversion and sensation seeking were unrelated to past-year alcohol use.

**Table 2.** Univariable Regression models for the predictors of interest

|  | *Dependent variable (SE):*<br>AUDIT-C |  |
| --- | --- | --- |
|  | Females | Males |
| *Personality traits*<sup>1</sup> |  |  |
| Positive urgency | 0.712<sup>***</sup> (0.089) | 0.952<sup>***</sup> (0.153) |
| Negative urgency | 0.680<sup>***</sup> (0.091) | 0.795<sup>***</sup> (0.156) |
| Lack of perseverance | 0.340<sup>***</sup> (0.087) | 0.552<sup>***</sup> (0.143) |
| Lack of premeditation | 0.403<sup>***</sup> (0.087) | 0.497<sup>***</sup> (0.153) |
| Sensation seeking | -0.020 (0.086) | 0.022 (0.151) |
| Conscientiousness | -0.385<sup>***</sup> (0.088) | -0.552<sup>***</sup> (0.141) |
| Extraversion | -0.072 (0.085) | 0.190 (0.141) |
| Neuroticism | 0.412<sup>***</sup> (0.094) | 0.546<sup>***</sup> (0.147) |
| Agreeableness | -0.241<sup>***</sup> (0.089) | -0.245<sup>*</sup> (0.140) |

*Note:* <sup>1</sup> All models were controlled for covariates (sociodemographic factors, genetic liability, and psychopathology). The table includes standardized estimates and standard errors for females and males separately. <sup>\*</sup> p<0.05; <sup>\*\*</sup> p<0.01; <sup>\*\*\*</sup> p<0.001

**Table 3.** Multivariable Regression

|  | *Dependent variable (SE):*<br>AUDIT-C |  |
| --- | --- | --- |
|  | **Females** | **Males** |
| Age | -0.301<sup>***</sup> (0.097) | -0.385<sup>**</sup> (0.154) |
| Race |  |  |
| Black vs. white | 0.821<sup>*</sup> (0.458) | -0.497 (0.692) |
| Other vs. white | -0.385 (0.377) | 0.479 (0.573) |
| Family history | -0.001 (0.088) | -0.062 (0.144) |
| Relationship status | 0.024 (0.173) | -0.893<sup>***</sup> (0.299) |
| Education |  |  |
| Bachelor vs. Graduate degree | -0.761<sup>***</sup> (0.264) | -0.514 (0.453) |
| Bachelor vs. High school or less | 0.093 (0.302) | 1.268<sup>**</sup> (0.525) |
| Bachelor vs. Post-secondary education | 0.106 (0.217) | 0.876<sup>**</sup> (0.377) |
| AUD severity | -0.079 (0.094) | -0.410<sup>**</sup> (0.165) |
| Major depression | -0.151 (0.095) | -0.177 (0.140) |
| Antisocial behaviors | -0.196<sup>*</sup> (0.104) | -0.417<sup>**</sup> (0.182) |
| Positive urgency | 0.462<sup>***</sup> (0.116) | 0.695<sup>***</sup> (0.204) |
| Negative urgency | 0.310<sup>**</sup> (0.124) | 0.272 (0.216) |
| Lack of perseverance | 0.100 (0.109) | 0.265 (0.199) |
| Lack of premeditation | 0.047 (0.109) | -0.235 (0.198) |
| Sensation seeking | -0.188<sup>**</sup> (0.094) | -0.063 (0.163) |
| Conscientiousness | -0.086 (0.111) | -0.282 (0.203) |
| Extraversion | -0.018 (0.089) | 0.243 (0.151) |
| Neuroticism | 0.082 (0.110) | 0.199 (0.186) |
| Agreeableness | -0.014 (0.097) | 0.033 (0.159) |
| Observations | 1,615 | 716 |
| R<sup>2</sup> | 0.074 | 0.140 |
| Adjusted R<sup>2</sup> | 0.063 | 0.115 |
| Residual Std. Error | 3.404 (df = 1594) | 3.846 (df = 695) |
| F Statistic | 6.389<sup>***</sup> (df = 20; 1594) | 5.638<sup>***</sup> (df = 20; 695) |

*Note:* The table includes standardized estimates and standard errors for females and males separately. \* \*\* \*\*\* p<0.01

In the multivariable models, considering all personality traits together while controlling for the covariates, results indicated associations between high positive urgency and greater past-year alcohol use in both sexes. In addition, we found that, in females, having greater negative urgency and lower sensation seeking were related to higher alcohol use. As for the roles of covariates, in both sexes, results showed associations between AUDIT-C scores and younger age, lower education, and lower antisocial behaviors. We note that the direction of the effect for antisocial behaviors was reversed in the multivariable models when we included personality traits related to impulsivity (see Figure S1 for correlations between the study variables). In males, being in a relationship and higher severity of AUD were also related to lower alcohol use. Finally, we observed differences in the variance explained by those models as a function of sex, with an adjusted R^2^ of 0.14 in males and 0.07 in females.

Though these analyses indicate sex-specific correlates of past-year alcohol use in individuals with severe AUD who attended AA, supplementary analyses did not reveal any sex-by-personality-trait interactions (Supplement).

## Discussion

This study aimed to investigate the personality correlates of alcohol consumption in people with severe AUD who have been involved in AA recovery groups. We explored the role of various personality and personality-like traits (namely extraversion, agreeableness, conscientiousness, neuroticism, negative and positive urgency, lack of premeditation, lack of perseverance, and sensation seeking) separately in males and females while controlling for a range of potential covariates.

First, we hypothesized that personality traits related to low perseverance/conscientiousness and high sensation seeking would be more related to higher alcohol use in males while emotional traits would be more important in females. Our results showed moderate associations between lack of perseverance/conscientiousness and AUDIT-C in the univariable models in both sexes, but this did not persist in the multivariable models. In males, in the fully-adjusted model, the only personality trait associated with past-year alcohol use was high positive urgency. Previous studies have underscored the role of low emotional stability (<u>Hakulinen and Jokela, 2019</u>) and positive urgency in heavy drinking among individuals with AUD (<u>Gowin et al., 2021</u>), but our results specifically indicate it in elevated alcohol consumption among adult males with severe AUD who attended AA recovery groups. In contrast, we did not find evidence that sensation seeking was related to alcohol use. Though sensation seeking may be an important correlate of excessive drinking (<u>Adan et al., 2017</u>), these findings suggest that it might not be a central factor in predicting recent drinking among those with severe AUD who pursued support through AA.

In females, findings underscore the influence of emotional traits, particularly positive and negative urgency, suggesting that the tendency to act rashly in emotional contexts might explain the risk of relapse in women with severe AUD who attended AA. We did not find an association between neuroticism and alcohol use in the multivariable model, but this could be explained by the influence of negative urgency. Indeed, previous studies reported that neuroticism was not directly associated with alcohol use but mainly acted through the effect of negative urgency (<u>Papachristou et al., 2016</u>). Results also implicate low sensation seeking in higher alcohol use among females. We observed the same pattern of results for antisocial behaviors in both sexes, suggesting that the effects of some of these externalizing characteristics might be confounded by other personality variables that remained associated with alcohol use in the fully-adjusted model. Beyond personality traits, our findings underscore the roles of additional variables.

Though prior results indicated that being single was a risk factor for heavy drinking after AUD treatment (<u>Sofin et al., 2017</u>), current results add a key nuance: being single represented an important risk factor for males but did not appear significant in females in either univariable or multivariable models. This difference is in line with well-established literature showing that (i) marriage is protective against heavy drinking in males (<u>Leonard and Rothbard, 1999</u>, <u>Umberson, 1992</u>) and (ii) divorce is causally related to increased risk of AUD with larger effect sizes in males than females (<u>Kendler et al., 2017</u>, <u>Salvatore et al., 2017</u>). This observation reinforces the need to conduct sex-specific investigations, as simply controlling for sex may hide the effect of important variables or wrongly implicate the significance of others. In addition, AUD severity was related to lower alcohol use in males who reported attendance to AA. Though this observation may appear inconsistent with a large body of studies showing that AUD severity is related to less favorable treatment outcomes (<u>Yoshimura et al., 2021</u>), the current sample is exclusively composed of individuals with severe AUD with a mean age of around 50 in males. In this sample, AUD severity might thus be related to negative consequences (<u>Andreasson et al., 2013</u>) that may increase the willingness to seek treatment, participate in self-help groups, and control their use of alcohol (<u>Kuramoto et al., 2011</u>).

Findings from this study should be understood in the context of some limitations. First, several factors may impact the representativeness of our sample: Participants were recruited from Facebook and ResearchMatch.org, thus individuals included in the study might not be representative of the whole population of individuals with AUD. While AUD is more common among males, the majority of our sample (65%) was female; this limitation is offset by our sex-specific analyses, which indicated no significant sex-by-personality interactions. In addition, our analyses focused on participants with complete data for all the measures of interest. Second, our assessment of AA consisted of a general question about lifetime participation without further details about the frequency or duration of AA involvement, and whether participation was voluntary versus court-mandated. It is also possible that participants attended other self-help groups. Future studies would benefit from more refined assessments of these factors, which could be included as covariates. Third, we relied on cross-sectional data, limiting our abilities to distinguish causes from consequences. Future studies should further investigate the longitudinal predictors of alcohol use in those with severe AUD who attended recovery groups. Finally, it was surprising to observe no association between family history of AUD and alcohol use in this sample of AA participants. However, a family history of alcohol problems was quite common in this sample: In a less severely affected sample, a wider range of variance in family history density could lead to a different result. It should be noted that, though measures of family history density have proved useful in previous studies (<u>Stephenson et al., 2022</u>), these measures do not allow a precise distinction between what part of the risk arises from genetic versus environmental transmission (e.g., the adversity related to having a parent with AUD). Preliminary evidence suggests that some genetic variants might be associated with medication-specific effects (<u>Biernacka et al., 2021</u>), but the role of environmental transmission in AUD behavioral treatment or self-help outcomes is still unclear. This question should be further explored by combining measures of aggregate genetic liability using both genotypic (polygenic risk scores) and phenotypic data.

The strengths of this study include our reliance on well-validated personality measures, sex-specific explorations, and analyses that control for a wide range of covariates that may not have been adequately accounted for in prior studies. Accordingly, the present study revealed distinct personality traits and sociodemographic factors associated with alcohol use in females and males with severe AUD who attended AA. Overall, results underscored the role of positive urgency in both sexes and the prominence of negative urgency in females only. Results also support an important role of age and education in both sexes as well as relationship status and AUD severity in males. These findings might help to better define who is at risk to continue drinking excessively after/during participation in AA. In particular, our results suggest that additional psychological interventions could be focused on potentially modifiable personality traits (<u>Roberts et al., 2017</u>): Interventions could target impulsive responses (particularly those associated with drinking) to positive and negative emotions. Interventions should also consider the influence of age and education and encourage social support for males who are not involved in a relationship.

## Data Availability

Data used in the present study are not currently available.

## Acknowledgments

This study was supported by the National Institute on Alcohol Abuse and Alcoholism (grant number: AA0026750). The use of REDcap was supported by an Award Number UM1TR004360 from the NCRR.

