## Supplement for "Personality correlates of past-year alcohol use in individuals with severe alcohol use disorder and a lifetime history of involvement in Alcoholics Anonymous"

**SUPPORTING INFORMATION**

**METHODS**

**Information about missing values**

There were 6% of missing data for age (N=183) and less than 1% for race (N=9), relationship status (N=9), education (N=8), and family history of alcohol problems (N=20). There were 7.5% of missing data for depression (N=222), and 1ess than 3% for impulsivity subcomponents: negative urgency (N=55), positive urgency (N=70), lack of perseverance (N=63), lack of premeditation (N=83), and sensation seeking (N=64). Finally, there was no missing data for antisocial behaviors or subcomponents of the Big 5 inventory.

**RESULTS**

**Descriptive analyses**

Figure S1. Depicts correlations between the study variables.

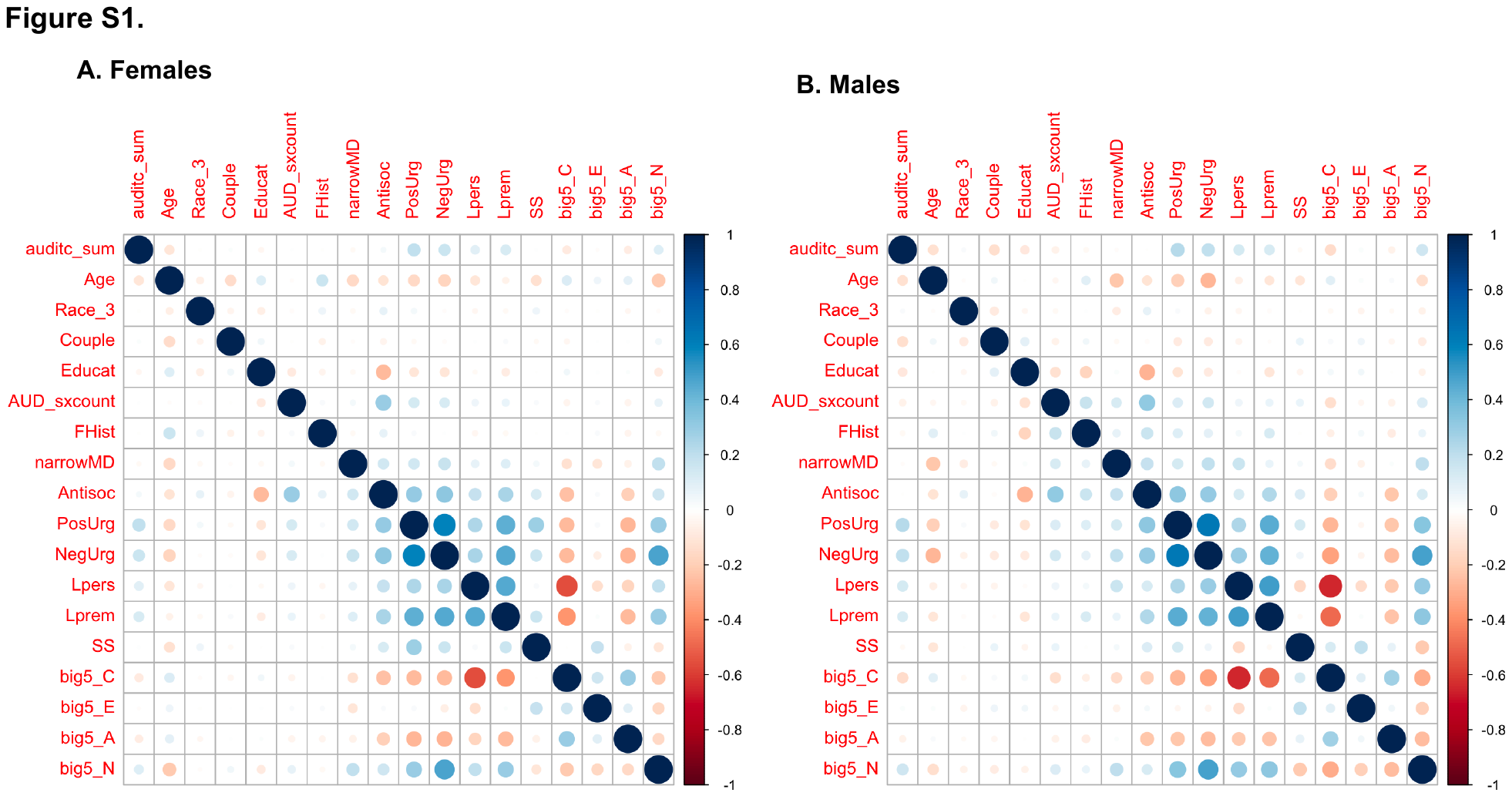

***Figure Caption***: auditc_sum = AUDIT-C score, Age = Current age, Race_3 = Race divided in 3 categories (White, Black, Other), Couple = Relationship status, Educat = Educational attainment, RecruitmentArm = Recruitment arm, AUD_sxcount = symptoms counts for Alcohol Use Disorder, FHist = Family History of alcohol problems, narrowMD = Major depression (DSM-V definition), Antisoc = Antisocial behaviors, PosUrg = Positive urgency, NegUrg = Negative urgency, Lpers = Lack of perseverance, Lprem = Lack of premeditation, SS = Sensation seeking, big5_C = Conscientiousness (big5), big5_E = Extraversion (big5), big5_A = agreeableness (big5), bug5_N = Neuroticism (big5). Pearson correlations were computed for two continuous variables, polychoric correlations for two categorical variables, and point biserial for one continuous and one ordinal variable.

**Linear regressions**

Univariable regression were performed between past-year alcohol use and each of our study covariates, results are described in Table S1.

| **Table S1.** Univariable Regression models for the study covariates | | |
| --- | --- | --- |
|  | Dependent variable:  AUDIT-C | |
|  | **Females** | **Males** |
| ***Covariates*** | | |
| Age | -0.461^***^ (0.085) | -0.646^***^ (0.130) |
| Race |  |  |
| Black vs. white | 0.856^**^ (0.405) | -0.415 (0.565) |
| Other vs. white | -0.067 (0.351) | 0.861^*^ (0.499) |
| Family history | -0.047 (0.082) | -0.075 (0.122) |
| Relationship status | 0.033 (0.160) | -1.073^***^ (0.255) |
| Education |  |  |
| Bachelor vs. Graduate degree | -0.890^***^ (0.243) | -0.832^**^ (0.387) |
| Bachelor vs. High school or less | 0.187 (0.273) | 1.432^***^ (0.426) |
| Bachelor vs. Post-secondary education | 0.072 (0.197) | 0.754^**^ (0.321) |
| AUD severity | -0.043 (0.084) | -0.209 (0.136) |
| Major depression | -0.011 (0.087) | 0.005 (0.125) |
| Antisocial behaviors | 0.139^*^ (0.081) | 0.292^**^ (0.130) |

**Interactions between sex and personality traits**

The main goal of this paper was to inform about the specific personality traits related to past-year alcohol use in females and males separately. We pursued additional analyses to test formal sex-by-personality-trait interactions. First, we ran a multivariable regression including sex, personality traits, and the study covariates. Results from this analysis supported the significance of sex (standardized beta = -0.72 [SE = 0.12], *p* < 0.001) and three specific traits: positive (standardized beta = 0.53 [SE = 0.10], *p* < 0.001) and negative (standardized beta = 0.31 [SE = 0.11], *p* = 0.004) urgency as well as sensation seeking (standardized beta = -0.16 [SE = 0.08], *p* = 0.04). This is in line with what we found in our sex-specific analyses. Second, we performed interaction analyses between sex and the three personality traits that reached significance in the multivariable model. We ran three analyses (one for each personality trait) including sex, personality, their interaction, and the study covariates. Results did not reveal any sex-by-personality-trait interactions at the 0.05 significance level (all betas < 0.19, all *ps* > 0.08).
